# Germline mutations and developmental mosaicism underlying *EGFR*-mutant lung cancer

**DOI:** 10.1101/2023.09.28.23296274

**Authors:** Risa Burr, Ignaty Leshchiner, Christina L Costantino, Martin Blohmer, Tilak Sundaresan, Justin Cha, Karsen Seeger, Sara Guay, Brian P Danysh, Ira Gore, Raquel A Jacobs, Kara Slowik, Filippo Utro, Kahn Rhrissorrakrai, Chaya Levovitz, Jaimie L Barth, Taronish Dubash, Brian Chirn, Laxmi Parida, Lecia V Sequist, Jochen K Lennerz, Mari Mino-Kenudson, Shyamala Maheswaran, Kamila Naxerova, Gad Getz, Daniel A Haber

## Abstract

While the development of multiple primary tumors in smokers with lung cancer can be attributed to carcinogen-induced field cancerization, the occurrence of multiple primary tumors in individuals with *EGFR*-mutant lung cancer who lack known environmental exposures remains unexplained. We identified ten patients with early-stage, resectable non-small cell lung cancer who presented with multiple anatomically distinct *EGFR*-mutant tumors. We analyzed the phylogenetic relationships among multiple tumors from each patient using whole exome sequencing (WES) and hypermutable poly-guanine (poly-G) repeat genotyping, as orthogonal methods for lineage tracing. In two patients, we identified germline *EGFR* variants, which confer moderately enhanced signaling when modeled *in vitro*. In four other patients, developmental mosaicism is supported by the poly-G lineage tracing and WES, indicating a common non-germline cell-of-origin. Thus, developmental mosaicism and germline variants define two distinct mechanisms of genetic predisposition to multiple *EGFR*-mutant primary tumors, with implications for understanding their etiology and clinical management.

## Introduction

As many as 10 percent of patients with NSCLC present with CT findings suggestive of two or more anatomically distinct synchronous lesions. This fraction is increasing with the overall increased utilization of CT imaging, and specifically with improved utilization of low dose CT screening in high-risk individuals with heavy smoking histories^1–4^. In such patients, multiple independent primary tumors are genetically unrelated, typically showing distinct genetic drivers, with mutational signatures of carcinogen-mediated DNA damage^1,5,6^. The concept of field cancerization in lung and other tissues, such as UV-exposed skin, explains the lifelong risk of multiple primary tumors and the need for regular cancer screening and monitoring in a subset of patients^1,5–13^.

NSCLC harboring activating mutations in the *EGFR* gene account for approximately 15% of all cases^14^. The canonical somatically acquired mutations strongly activate receptor signaling, driving tumorigenesis and leading to dramatic clinical responses to EGFR kinase inhibitors^15,16^. Canonical *EGFR* mutations do not show smoking-associated mutational signatures, and the dramatic enrichment of cases among never smokers with NSCLC (up to 50% of cases) indicates that these mutations are linked to other risk factors^17,18^. Remarkably, the incidence of *EGFR*-mutant NSCLC is almost twice as high in women, compared with men, and in Asian populations, compared with non-Asian populations. While germline genetic polymorphisms linked to *EGFR*-mutant NSCLC have not been identified, shared haplotypes have been described between Asian and South American populations at risk of *EGFR*-mutant cancer^19–22^.

Canonical somatic activating *EGFR* mutations (e.g., L858R) have never been observed in the germline, suggesting that their cellular signaling activity is incompatible with normal embryonic development. However, we previously identified a family with inherited susceptibility to *EGFR*-mutant lung cancer, caused by a germline variant with attenuated signaling activity^23^, the T790M “gatekeeper” mutation commonly associated with acquired drug resistance to first and second generation EGFR inhibitors^24^. In this family, inheritance of a germline T790M-*EGFR* mutation confers weakly enhanced EGF signaling, which may be tolerated during lung development, but is then followed by the development of multiple tumors, each having a somatic canonical *EGFR* mutation *in cis* with the inherited variant, which has been shown to synergize and confer enhanced activated signaling^25^. While extraordinarily rare, familial susceptibility to NSCLC due to an inherited *EGFR* T790M allele has since been confirmed in a few additional families^26,27^ and other rare *EGFR* germline familial variants, including V843I, R776X, and P848L, have been reported^28^.

In the absence of smoking-associated field cancerization or known familial predisposition, the presence of multiple synchronous *EGFR*-mutant tumors appears paradoxical, and several distinct models have been proposed. Deep sequencing of normal lung tissues has revealed rare oncogenic *EGFR* mutant alleles in 18% of samples^29^, consistent with the emerging appreciation that cancer-causing mutations may populate apparently healthy aging tissues^30–34^. Any tumors ultimately derived from such mutant *EGFR*-harboring cells would constitute independent genetic events. On the other hand, previous studies of multiple primary *EGFR*-mutant lung cancers have indicated the presence of shared mutations, leading to the suggestion that they may be clonally related metastases, potentially resulting from intra-pulmonary spread through lymphatics and possibly even airspaces, in the absence of disseminated metastatic disease ^5,6,9–13^. However, such localized intra-pulmonary mechanisms of dissemination do not readily explain involvement of different lobes and contralateral lungs, which is commonly observed in such cases. Moreover, many lung lesions in patients with *EGFR*-mutant multiple primary tumors are histologically pre-invasive, without evidence of lymphovascular or visceral pleural invasion, thereby reducing the likelihood of metastatic spread. Given these considerations, we sought to reexamine these conceptual models and test whether other genetic mechanisms may explain *EGFR*-mutant lung cancers presenting with multiple primary lesions.

## Results

### Patient Clinical Characteristics

We identified ten patients from medical records at Massachusetts General Hospital who had surgery between 2004-2019 for multiple early-stage, spatially distinct lung adenocarcinomas, with at least one specimen positive for *EGFR* mutation by routine clinical genotyping (Table 1, patients 1-10). No patient had received any treatment prior to surgery, and none were found to have lymph node involvement or suspected metastatic disease. Four patients were never smokers, three patients had a remote smoking history of <5 pack years, and three patients had a former >30 pack year smoking history. None of the ten patients had a family history that was considered remarkable for multiple malignancies, including lung cancer. Tumor diameters ranged from 0.5 cm to 3 cm. In five patients, multiple tumors involved bilateral lungs, while in three patients, tumors arose within different lobes on the ipsilateral side and in two they were confined to a single lobe (Table 1, Supplementary Table S1 and Supplementary Figure 1a). Histologically, they were classified as precancerous atypical adenomatous hyperplasia (AAH, 2 tumors from 2 cases), adenocarcinoma-in-situ (AIS, 1 case), minimally invasive adenocarcinoma (MIA, 10 tumors from 6 cases), mixed AIS and MIA (1 case), and invasive adenocarcinoma (18 tumors from 8 cases). In addition to the above cases of sporadic lung cancer, we applied our molecular analyses to a family with known germline transmission of an *EGFR* T790M allele (noncritical clinical features in the family history have been changed to preserve confidentiality) (Table 1; Figure 1a, Supplementary Figure 1b) ^23^. In this family, the number of tumors per individual mutation carrier ranged from 1 to 13, with histology ranging from AIS to invasive adenocarcinoma (Figure 1b-c). We first validated our tumor molecular analyses in one patient from this family with known germline susceptibility, and then applied the same analytics to the sporadic cases with multiple primary tumors.

**Table 1:** Demographic and Clinical Characteristics of the Patients. Clinical characteristics of two patients from familial lung cancer pedigree (III-1, III-4) and 10 patients with sporadic multiple primary lung cancers. Stage refers to the clinical impression at the time of surgical resection following IASCLC 8th edition staging guidelines. Number of tumors corresponds to those observed on CT throughout patient’s lifetime. Additional clinical details for each patient are found in the Supplementary Information.

| Patient | Age Range;<br>Gender | Smoking | Genetic<br>Ancestry | Stage | # Tumors | Contralateral | Multilobar |
| --- | --- | --- | --- | --- | --- | --- | --- |
| III-1 | 51-55 M | moderate | European | 0 | >13 | Y | Y |
| III-4 | 61-65 F | never | European | 0 | 10 | Y | Y |
| 1 | 61-65 F | former | European | IA | 4 | N | Y |
| 2 | 81-85 M | former | European | IA | 3* | N | N |
| 3 | 51-55 F | never | African | IA | >3 | N | Y |
| 4 | 56-70 F | former | Asian | IA | >5 | Y | Y |
| 5 | 51-55 F | never | African | IA | 5 | Y | Y |
| 6 | 71-75 F | former | European | IIB | 4 | N | N |
| 7 | 76-80 F | never | European | IB | 2 | N | Y |
| 8 | 81-85 F | former | European | IA | 4 | Y | Y |
| 9 | 61-65 F | never | European | I | 4 | Y | Y |
| 10 | 66-70 F | former | European | IA | 4 | Y | Y |

\* additional tumors observed upon pathological assessment

**Figure 1.**
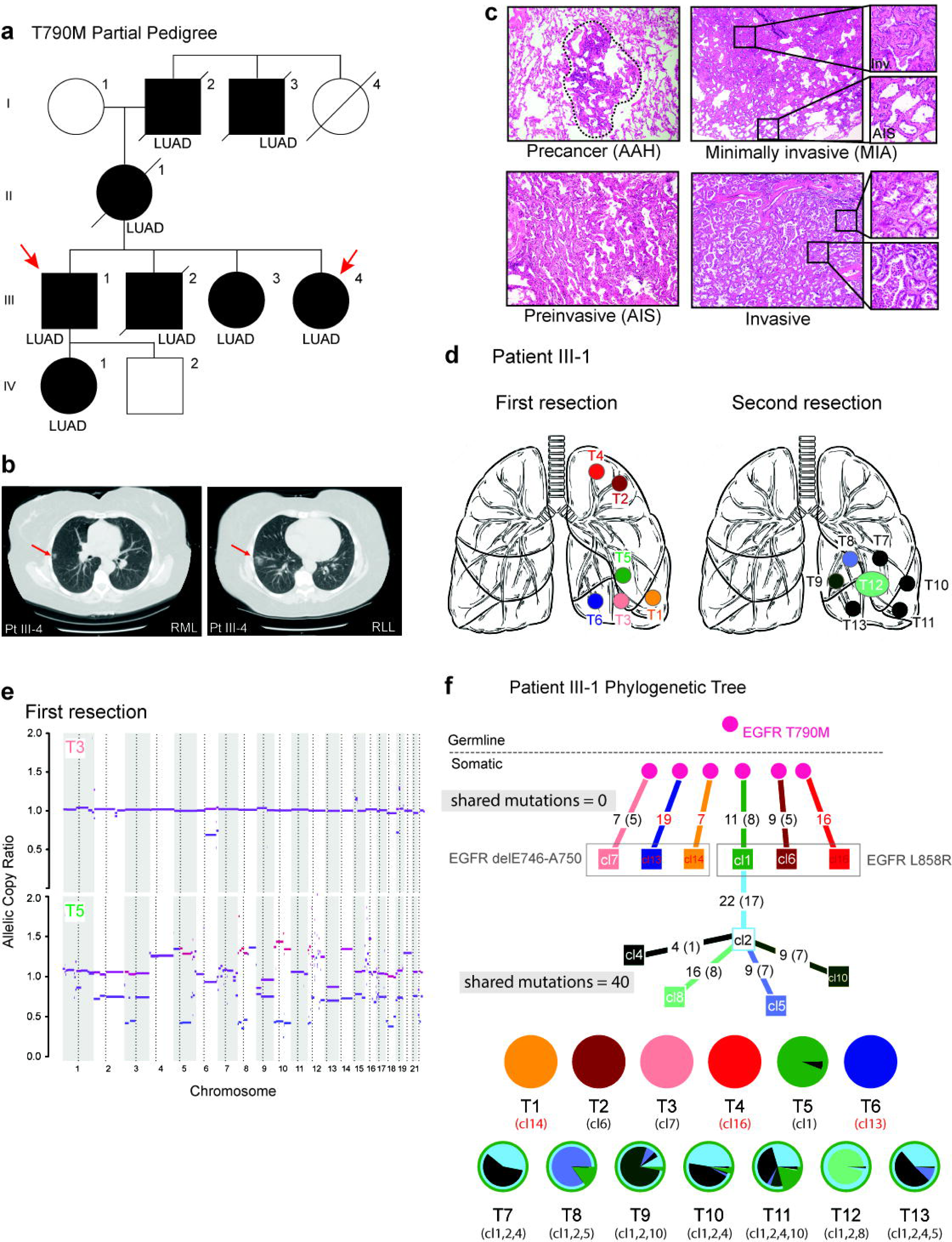
Genetic analysis of familial lung cancer due to inherited T790M mutation in the *EGFR* gene. **a,** Partial pedigree of a family with multiple cases of lung adenocarcinoma, in which the index case (III-1) was diagnosed with six primary carcinomas (first resection), followed by resection of seven tumors ten years later. These tumors were available for analysis as were three tumors from his sibling (III-4). Individuals shown in black have a confirmed or obligate germline *T790M-EGFR* mutation and those who have developed lung adenocarcinoma are denoted (LUAD). The pedigree was minimally altered to preserve confidentiality (males, square; females, circles). **b,** Computed tomography scans of two tumors from patient III-4, one in the Right Middle Lobe (RML), and the other in the Right Lower Lobe (RLL). **c,** Histology of tumors from *T790M-EGFR* family patients showing the range of invasiveness encountered in our cohort, from precancerous Atypical adenomatous hyperplasia (AAH; patient III-4 lesion T2), to Adenocarcinoma in situ (AIS; patient III-4 lesion T2), to minimally invasive adenocarcinoma (MIA; patient III-1 lesion T2), to invasive adenocarcinoma (patient III-1; lesion T12). Panels are at 40x magnification, with insets at 200x. **d,** Schematic of the tumor locations in patient III-1, at the first resection (left) and the second resection 10 years later (right). **e,** Copy number data for two tumors from the first resection of patient III-1. Tumor T3 is a MIA and shows an allelic copy ratio of 1.0 for most chromosomes, indicating diploidy. Tumor T5 is an invasive adenocarcinoma and shows extensive aneuploidy. **f,** Phylogenetic lineage tracing of multiple tumors from patient III-1 based on Whole Exome Sequencing with PhylogicNDT. Theoretical cell populations are circles and clones derived from the WES are squares (e.g. c1). The germline *EGFR* mutation found in normal lung tissue is denoted at the top. The branches are configured based on shared and distinct mutations in each clone. Numbers within lineage tracings represent the number of new additional exomic mutations identified in each clone. The common somatically acquired *EGFR* mutations are shown, with the clones where they were identified in grey boxes. We cannot determine whether clones that share a boxed EGFR mutation developed independently or from a shared precursor. Resected tumors are assigned to clones based on their majority population in the pie charts shown below the tree. The tumors from the first resection, T1-T6, share no mutations outside of EGFR, as represented on the tree by no intersection point for clones 1 and 12-16, and in the pie charts by no colors shared between pie charts. In contrast, the tumors from the second resection, T7-T13, share 26 mutations, as represented by the long trunk leading from cl1 to cl2 before branching into cl4, 5, 8, and 10. Additionally, the pie charts for these tumors are complex mixtures of these four clones and clones are shared among multiple tumors.

### Molecular evolution in familial *EGFR*-mutant lung cancer

Multiple specimens were available for two patients in the family with an inherited T790M mutant *EGFR* allele. For the index case (patient III-1), six geographically distinct lesions were resected from the left upper and lower lobes at the time of initial diagnosis, and an additional seven lesions were resected ten years later from the left lower lobe (Figure 1d). Whole exome sequencing (WES) of macrodissected paraffin-embedded tumor sections, compared with normal blood specimens, confirmed the heterozygous T790M *EGFR* germline mutation in all tissues. Two representative independent tumors from the first resection are shown in Figure 1e, including T5, which has increased CNA and subsequently gave rise to metastatic disease. All tumors shared the functionally attenuated T790M germline mutation, and they showed subsequent somatic acquisition of a single secondary canonical *EGFR* mutation, either L858R or an exon 19 deletion mutation. However, beyond *EGFR*, the initially resected six tumors have no shared somatic mutations, consistent with their independent origin in the setting of cancer predisposition (Figure 1f).

In contrast, exome sequencing and phylogenetic reconstruction of the seven tumors resected ten years later (T7-T13) shows them to share between 15 and 41 somatic mutations: (average 29.8% of all mutations are shared between the initial lesion T5 and the later lesions T7-13, and average 81.4% of all mutations are shared across the later lesions T7-13 (Figure 1f, Supplementary Table S3). Published analyses of intratumoral heterogeneity in lung adenocarcinomas indicates that any two regions of the same tumor share approximately 70% (interquartile range 50-80%) of all mutations detected by WES^35^. Using this as a benchmark for our analysis, we conclude that the later tumors were recurrent metastatic foci, derived from one of the originally resected tumors (T5). This proof-of-concept analysis illustrates the genetic parameters that define completely independent early lung tumors *versus* metastatic recurrences from a single primary tumor, all arising within the context of an inherited genetic susceptibility.

### Novel germline EGFR variants in sporadic cases with multiple primary tumors

Having distinguished independent primary tumors from metastatic recurrences in the setting of familial *EGFR*-mutant lung cancer, we turned to a separate cohort of ten apparently sporadic cases with multiple *EGFR*-mutant lesions. In two cases, we identified uncommon heterozygous germline *EGFR* variants in normal lung tissue. The lung tumors showed the same heterozygous mutation, along with a second, somatically acquired canonical *EGFR* mutation. The germline *EGFR* variants are within critical functional domains of the protein: patient 4 has a G873E mutation within the tyrosine kinase domain (exon 21) and patient 5 has a H988P mutation within the autophosphorylation domain of EGFR (exon 25) (Figure 2a). Both of these patients had bilateral synchronous lesions at presentation (Figure 2b). WES identified no shared somatic mutations, outside of the *EGFR* gene, indicating independent primary tumors arising in the setting of potential genetic predisposition, analogous to the first six tumors characterized in index patient III-1 from the prototype T790M-*EGFR* family (Supplementary Table S2, Figure 2c). When present in the germline, *T790M*-EGFR is a weakly activating allele, and lung cancers show somatic acquisition of a canonical *EGFR* activating mutation *in cis* with the germline allele, resulting in a strongly activating protein with both mutated residues^23,25^. In patient 4, the secondary canonical mutation L858R was also found *in cis* with the germline allele in all three tumors; patient 5 also had secondary (L858R, exon19 delE746-A750) alleles, but given the position of the relative mutations, the length of sequencing reads precluded determination of whether the heterozygous secondary mutation arose in *cis* or in *trans* with the heterozygous germline allele (Supplementary Figure 2a and Supplementary Table S1, S2).

**Figure 2.**
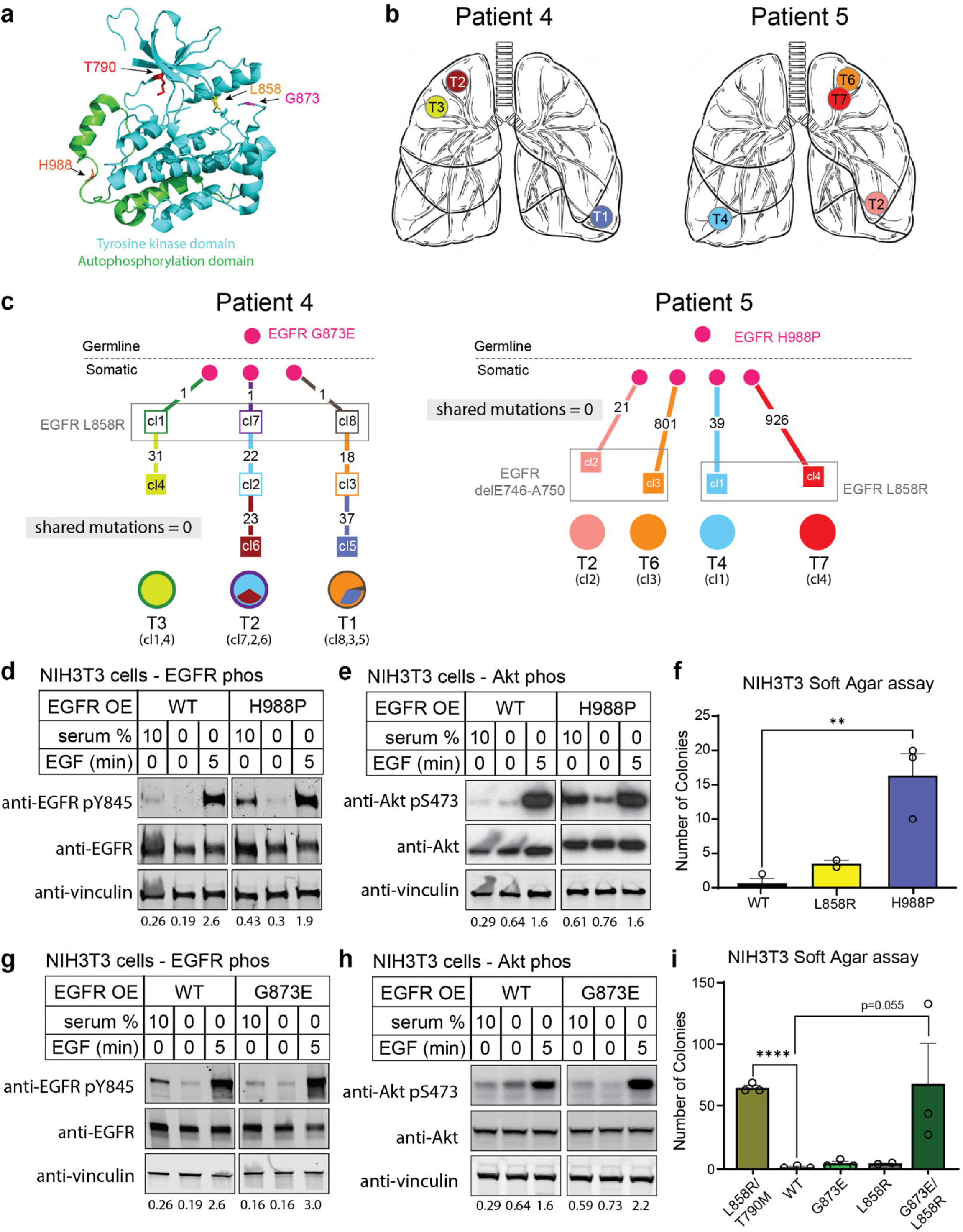
Germline H988P and G873E mutations increase EGFR activity. **a,** Position of the H988P, G873E, L858R, and T790M mutations within a partial EGFR protein crystal structure (aligned PDB structures EGFR 696-1022 T790M (5gty) and EGFR 703-985 (4zjv). H988 in orange, G873 in magenta). The autophosphorylation domain (shown in green) is adjacent to the catalytic tyrosine kinase domain (shown in blue). S1060 is not in this structure as it only exists in a different isoform. The H988P mutation is within the autophosphorylation domain and the G873E mutation is in the tyrosine kinase domain. **b,** Schematic of the multiple tumor locations in patients 4 and 5. **c,** Lineage tracing of the three tumors in patient 4 and the four tumors in patient 5, derived from Whole Exome Sequencing (WES). The constitutional *EGFR* mutation found in normal lung tissue and in all tumors is denoted at the top of the trees. Somatically acquired canonical activating *EGFR* mutations occurred in all tumors. We cannot determine whether clones that share a boxed EGFR mutation developed independently or from a shared precursor. Pie charts below the tree indicate clonal representation within each resected tumor. As with the first resection for patient III-1 (Figure 1f), these trees show no intersection between the branches, indicating independent tumors that only share the *EGFR* mutation. Consistently, the pie charts do not share any color between tumors. **d-e,** Functional effect of the H988P *EGFR* mutant, compared with wild type construct, assayed using western blotting of EGFR phosphorylation (D) or downstream Akt signaling (E) as markers of EGFR activation in mouse NIH 3T3 cells, which lack endogenous *EGFR* expression. EGFR phosphorylation at tyrosine 845 (Y845) and Akt phosphorylation at serine 473 (S473) are measured, quantified by total EGFR protein control and vinculin loading control, under three culture conditions: baseline culture (10% fetal bovine serum), serum starvation (24 hr) and 5 min after addition of EGF (100 ng/mL) to starved cultures. The H988P mutant shows constitutive signaling, compared with wild type protein under baseline unstimulated conditions. All blots within a subfigure came from the same gel. Quantifications below the blots correspond to the normalized average p-EGFR signal as detailed in Supplementary Figure 3. **f,** Quantification of colony formation by NIH 3T3 cells in soft agar, a correlate of tumorigenic potential, in cells expressing either wild type, L858R mutant, or H988P mutant *EGFR* constructs. Quantitation represents three independent experiments. Error bars are SEM. P-values are a one-tailed unpaired t-test. **p<0.01. **g-i,** EGFR and Akt phosphorylation and quantification of soft agar colony formation in cells expressing G873E, analyzed as in Fig 2f. ****p<0.0001 The G873E mutation itself does not alter EGFR signaling measurably, but it synergizes with an in cis canonical L858R mutation. The in cis L858R/T790M mutant allele is shown as a control.

The *EGFR* mutation H988P has been previously identified as a variant of unknown significance^36^. Therefore, we tested its functional properties in reconstruction experiments using standard signaling assays in mouse NIH/3T3 cells which do not express the endogenous protein. Compared with wild-type *EGFR,* the H988P mutant shows modestly increased phosphorylation under unstimulated conditions, a measure of baseline receptor signaling activity, together with enhanced downstream signaling of its key mediator AKT serine/threonine kinase (Figure 2d-e, Supplementary Figure 3a-b). *H988P-EGFR* transfected NIH/3T3 cells also generate more colonies in soft agar, a prototype cell transformation assay (Figure 2f, Supplementary Figure 3c). The second germline *EGFR* variant, G873E, has been reported as a somatic mutation and shown to play a role in resistance to gefitinib^37–40^. By itself, we find that it has modest activating capacity, but it is synergistic when combined *in cis* with the canonical L858R mutation (Figure 2g-i, Supplementary Figure 3c-e). Importantly, all tumors from patient 4 had L858R mutation *in cis* with the germline G873E mutation (Supplementary Figure 2a). Thus, like the established familial T790M mutation, both H988P and G873E appear to have an attenuated proliferative effect that may be tolerated in the germline without compromising normal embryonic development, but the inherited alleles subsequently sustain additional somatic *EGFR* mutations ultimately leading to malignancy.

Two other cases arising in minimal smokers harbored tumors that were genetically independent. In one case, patient 6, we identified a germline *EGFR* S1060A mutation, residing within the alternative splicing variant *EGFR-vA*, which is expressed at low levels in normal tissues (Supplementary Figure 2b). The *EGFR-vA* isoform has been reported as potentially oncogenic in gliomas^41^, but we were unable to confirm aberrant EGFR signaling associated with S1060A, making it a variant of unknown significance. In the other case, patient 1, we did not identify a candidate functional germline mutation that might explain the independent somatic genetic origin of these tumors (Supplementary Figure 2c).

### Developmental Mosaicism in Multifocal Lung Cancer

Among the remaining six cases with multiple primary *EGFR*-mutant cancers, we identified metastatic disease in two cases, patients 2 and 3; in both, recurrent disease was considered among the clinical possibilities at the time of resection. In these cases, WES and phylogenetic reconstruction (Methods) showed that many mutations were shared across all tumors (16 and 37 shared mutations in patients 2 and 3, respectively, representing an average of 36.6% of all exonic mutations within a tumor (range from 6.8% to 100% per tumor) (Figure 3, Supplementary Tables S2, S3). This is consistent with the published range of 50-80% clonal mutations between two samples of the same tumor, and with our findings in the T790M family^35^. Both cases subsequently had clinically recurrent cancer within 3 years of surgery.

**Figure 3.**
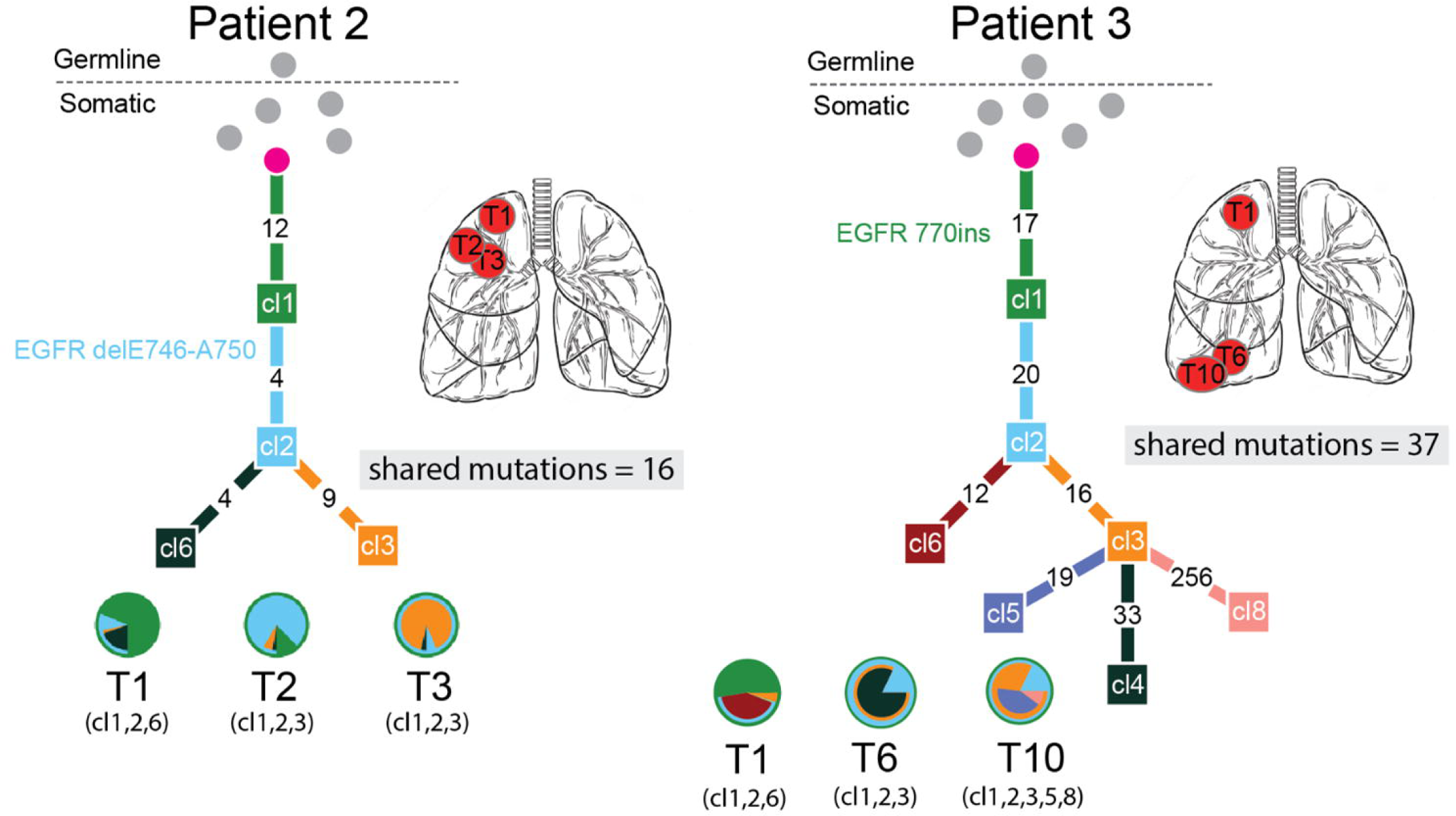
Lineage tracing of metastatic cancers. Phylogenetic trees from WES of two metastatic patients. Numbers on branches are mutations that accumulated between two nodes. As with the second resection for patient III-1 (Figure 1F), these trees show a long trunk of shared mutations before branching into separate clones. Consistently, the pie charts are made up of mixtures of clones that are shared between tumors.

For the remaining four cases (patients 7-10), WES revealed a moderate number of shared somatically acquired mutations across anatomically separate tumors (range 1 to 6 shared mutations, representing 0.2% to 5.3% (average 1.4%) of all exonic mutations) (Figure 4). This unusual pattern argues against completely independent tumors arising in the context of germline genetic predisposition, for which we would expect no shared mutations, other than *EGFR*, across anatomically distant tumors. It is also readily distinguishable from clonally-related metastatic lesions, based on the relatively low fraction of shared somatic mutations. Notably, *EGFR* mutations themselves were identical across anatomically distinct tumors within individual patients: in one case (patient 7), geographically distinct tumors shared a very rare mutation (SPKANKEI752del) that is unlikely to have developed twice independently, and in another case (patient 8), two tumors shared five unique somatic mutations, in addition to the common canonical *EGFR* L858R mutation (Figure 4a-b, Supplementary Tables S2, S3). These data are suggestive of mosaicism: distant shared somatic ancestry among multiple primary tumors within individual patients.

**Figure 4.**
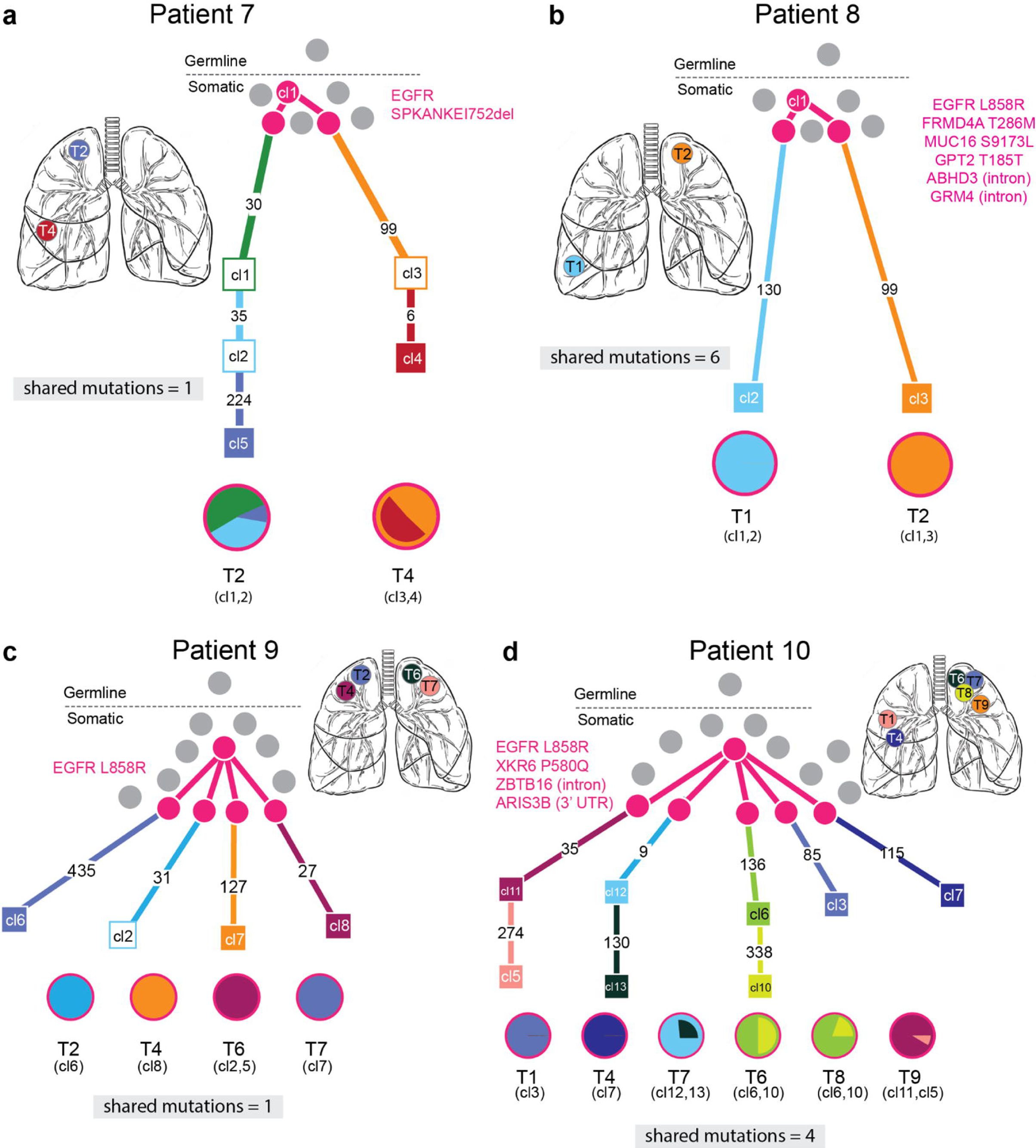
Mosaic somatic *EGFR* mutations mediate multiple primary lung tumors resulting from shared common ancestors. WES-derived phylogenetic trees of four cases with multiple tumors that share a common somatic ancestor. The shared somatic mutations, including *EGFR*, are shown in magenta. Numbers on branches are mutations that accumulated between two nodes, which represent distinct clones identified by WES. In comparison with the previous cases, the branches of these trees do intersect at the pink clones, indicating some shared genetic ancestry that is not observed in the completely independent or germline tumors. However, the number of shared mutations and the trunk of shared ancestry is very small relative to the total number of mutations in each clone. This is distinct from the patients with metastatic cancer. The pie charts of these tumors all exhibit the pink clone, but are otherwise relatively simple and do not share clones between tumors. **a,** In case 7, the two geographically distinct tumors share an extremely rare somatic *EGFR* mutation, SPKANTKEI752del, and then acquire 105 and 289 separate exomic mutations. **b**, In case 8, both tumors share the recurrent mutation L858R, in addition to five somatic mutations, before acquiring 105 and 136 separate mutations each. **c,** In case 9, four tumors share the L858R mutation before acquiring between 27 and 435 private mutations. **d,** In case 10, six tumors share L588R, in addition to three somatic mutations, before acquiring 85-474 private mutations.

Mosaicism in adult tissues predicts that variant alleles may be present, albeit at very low frequency, within normal tissues, where they may comprise a reservoir of cells susceptible to transformation^42^. To detect such cells, we employed droplet digital PCR (ddPCR) technology, which counts individual DNA molecules with a detection limit of ≤0.01% of cells within a population^43^. In all three putative mosaic cases with L858R*-EGFR* mutant tumors, the L858R mutation was detectable in multiple anatomically distinct normal lung samples, albeit at much lower allele fractions than in the tumor samples (Figure 5a). As a control, no such mutations were detected within normal tissues of cases with *EGFR* mutations other than L858R (patients 1 and 7). These molecular analyses suggest the presence of very rare cells harboring shared *EGFR* mutations within normal lung tissues of some patients with potentially mosaic-derived *EGFR* mutant cancers.

**Figure 5.**
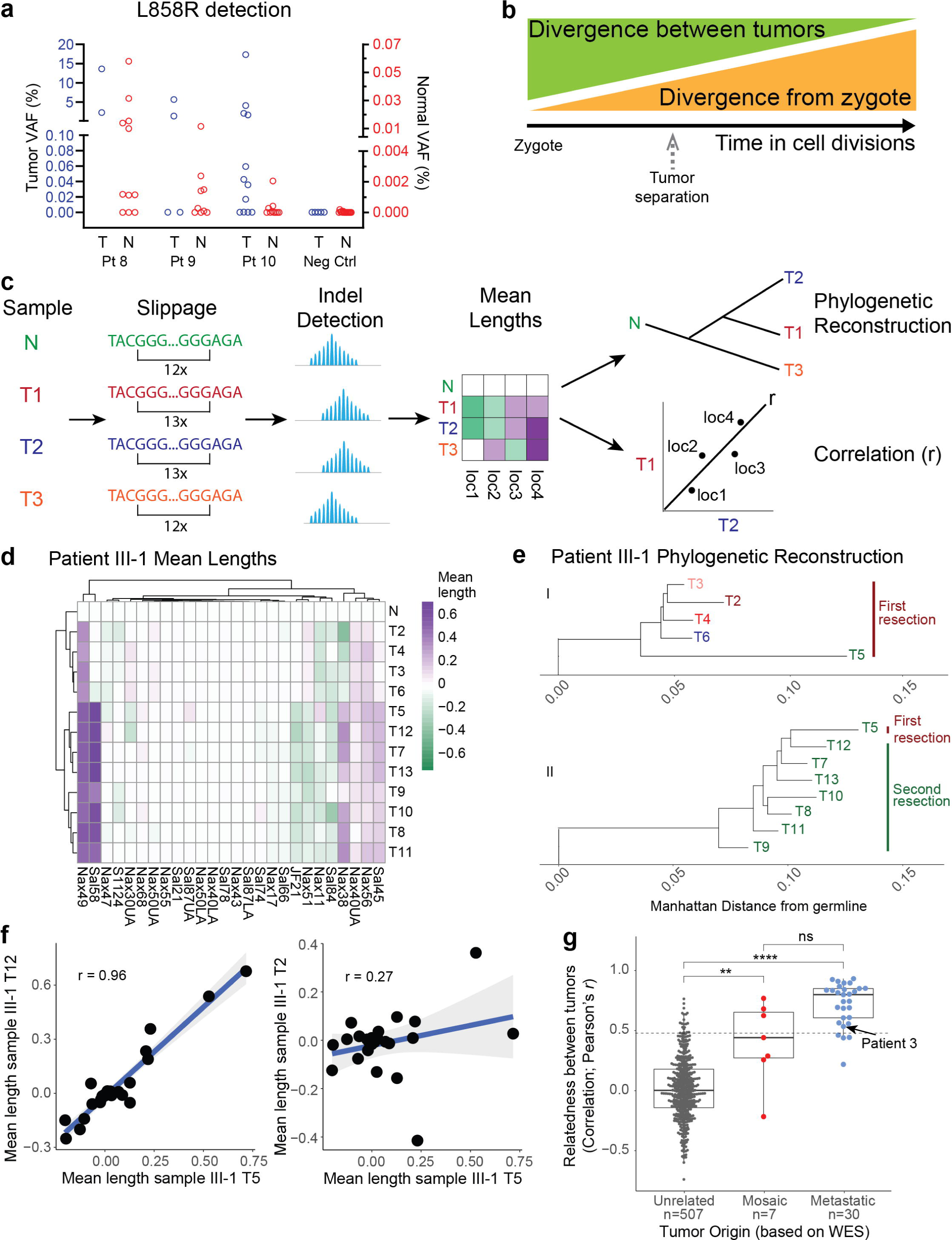
Developmental mosaicism is demonstrated by mutated normal lung cells and early common ancestors. **a,** Detection of the L858R *EGFR* mutation using droplet digital PCR (ddPCR) within normal lung tissue, in three cases where the tumor harbors the same mutation, but not in cases where the tumor contains another *EGFR* mutation (Neg Ctrl, patients 1 and 7). The variant allele frequency (VAF, EGFR L858R mutant copies/total EGFR copies) for tumors is on the left y-axis (blue) while the lower VAF in normal samples is on the right y-axis (red). **b,** A conceptualization of how poly-G correlation analysis reflects the relative time at which two related tumors diverged from each other and from the normal tissue. As the point of tumor separation (e.g. spontaneous initiation or metastasis) moves away from the zygote in time (divergence from zygote), the number of cell divisions that may have occurred after the tumors separated from each other decreases (divergence between tumors). The divergence between tumors is reflected in the poly-G analysis as the inverse of the correlation (*r*). This model shows that as *r* increases (1/*r* decreases), the relative time of divergence moves closer to the present. **c,** Schematic of poly-G genotype analysis method, based on Naxerova et al, 2017^45^. Tumor and normal samples collected from a single patient have poly-G sites that may have undergone slippage due to hypermutability. The PCR-based assay detects these indels and measures their mean length change compared to normal tissue. These changes compared to normal are represented in heat maps. Comparing two tumors produces a single point for each poly-G site. The correlation between the two tumors across all poly-G sites is represented by the Pearson’s correlation coefficient (*r*). **d,** Representative heatmap showing the mean distance from normal lung for each poly-G hypermutable region, for each tumor from patient III-1. The stronger the color, the more different the region is from normal. Tumors that have very similar patterns are more closely related than tumors that have different patterns. **e,** Phylogenetic tree of patient III-1 based on the poly-G derived Manhattan distance reconstructed using the neighbor joining method. The tree is rooted at the germline sample. A vertical bar at the leaves of the phylogenetic tree shows the resection time of each tumor sample. I) shows only samples from the first resection and II) shows samples from the second resection plus recurring sample T5. **f,** Representative correlation plots between tumors from patient III-1 showing how Pearson’s r measure of relatedness was determined. Two tumors being compared are on each axis (T12 and T5 in left graph and T2 and T5 in right graph) and the dots represent the mean length from normal at each poly-G location. *r* estimates what fraction of cell divisions were shared between the tumor pair before they diverged. The grey shading represents the 95% confidence interval of the correlation. **g,** Relatedness between tumors within a patient or between patients, as quantified by the poly-G correlation between tumor pairs. Each point represents the poly-G evolutionary distance between two tumors from cases that are unrelated (different individuals), mosaic or metastatic (classification based on WES sequencing analysis of exonic mutations). Unrelated tumors have fewer shared divisions than either mosaic or metastatic tumors. The dotted line represents that correlation coefficient below which 95% of the unrelated tumor pairs fall. Box plot elements: center line, median; box limits, lower and upper quartiles; whiskers, lowest and highest value within 1.5 IQR. p-values are a Holm-Bonferroni corrected post-hoc Dunn’s test after a significant Kruskal-Wallis test. **p=8.5E-3, ****p=6.4E-18. The arrow is the tumor pair from metastatic patient 3.

To quantify the divergence between tumors, we generated poly-G fingerprints^44–46^, which measure insertions/deletions in hypermutable guanine mononucleotide repeats. These mutations occur at high rates during DNA replication as a consequence of polymerase slippage^47^ (Figure 5c). Therefore, the divergence between the poly-G genotypes of two somatic cell populations is a reflection of the number of cell divisions that separate them. We first benchmarked the poly-G assay on samples from *EGFR*-T790M familial cancer patient III-1. Poly-G analysis demonstrated a short shared evolutionary history and few shared variants among the tumors from the first resection, consistent with the results from the WES analysis. In contrast, the metastatic lesions from the second resection 10 years later showed a long shared trajectory and close genetic concordance (Figure 5d-e). In accordance with the WES results, poly-G analysis conclusively showed that tumor T5 from the original resection was the source of metastatic disease.

To enable a quantitative comparison of the genetic relatedness between any pair of tumors from the same patient, we calculated Pearson’s correlation coefficients among the tumors’ poly-G genotypes (Supplementary Table S4). The correlation coefficient estimates the fraction of cell divisions in the history of two tumors that they have spent as part of the same lineage. Crucially, this estimation does not depend on knowledge of the underlying mutation rate or purity of the tumors, thereby providing an unbiased view of their evolutionary history. Thus, tumor samples that share a large fraction of their evolutionary history (such as metastases) show high correlations (Figure 5f, left panel), while tumors that share limited evolutionary history display low correlations (Figure 5f, right panel). To determine the evolutionary history that two unrelated tumors would be expected to share by chance, we calculated the correlation between all tumor pairs from different patients (Figure 5f, unrelated group). Indeed, we found the average correlation of these unrelated tumors was 0.02, with the 95^th^ percentile being 0.46 (Figure 5f, dotted line). The analyzed tumors from patient 3, categorized as metastatic based on exonic mutations, displayed a correlation of 0.53 and thus exceeded the 95^th^ percentile of unrelated tumors (Figure 5f, arrow). This means that they underwent 53% of their cell divisions as part of the same lineage and is consistent with the large number of shared exonic mutations.

Poly-G analysis of patients 7-10, with potential mosaically-derived multiple primary tumors, supports the notion that the lineages giving rise to these cancers diverged relatively early in time (Supplementary Table S3). On average, the tumors in these patients are less closely related than *bona fide* metastases, but they share a longer developmental history than completely independent tumors. We calculate that the lineages giving rise to these tumors underwent on average 44% of their cell divisions together before separating, compared with 80% for metastases and 0% for unrelated tumors (Figure 5g). Remarkably, three of the four patients with potential mosaically-derived primaries had tumors located in contralateral lungs, and one had tumors located in two different lobes of the same lung (Supplementary Figure 1), yet histopathological analysis in all four patients shows only four of 14 tumors to be invasive adenocarcinomas, with the other 9 lesions being minimally invasive (Supplementary Table S1). Thus, both genetic data and histopathology suggest that metastasis is unlikely to explain the appearance of multiple primary tumors in these cases. Consistent with the sequencing and poly-G lineage studies, the fact that distinct tumors commonly arose in contralateral lungs suggests that their divergence may have occurred early in lung development, with their progeny seeding disparate regions of the adult organ.

## Discussion

We have shown that multiple primary *EGFR*-mutant lung cancers may arise from an unappreciated attenuated germline variant in the *EGFR* gene or from canonical activating *EGFR* mutations that are acquired early in development leading to mosaicism for this driver mutation in the adult lung (Figure 6). The multiple primary tumors arising in the setting of germline predisposition share no somatic mutations, other than primary germline and secondary somatic *EGFR* mutations, and poly-G analysis shows a limited shared evolutionary history among them. In contrast, the multiple primary tumors that emerge in the absence of a germline *EGFR* mutation share identical somatic canonical *EGFR* mutations along with mutations in other genes (0.2 to 5.3% of all exonic mutations), confirming their shared clonal origin and suggestive of developmental mosaicism. Poly-G lineage analysis indicates that such mosaically-derived tumors display common ancestry that is intermediate between the short shared lineage seen in independent tumors with germline predisposition and the longer shared lineage displayed by metastatic tumors. Metastatic lesions are also readily distinguished from mosaically-derived primary tumors by their much higher fraction of shared mutations among each other and with their primary tumor of origin (50-80% of all exonic mutations). Thus, the appearance of multiple primary *EGFR*-mutant tumors is biologically distinct from the metastatic spread of cancer, and it may result from an initiating *EGFR* mutation, either in the germline or during early development.

**Figure 6.**
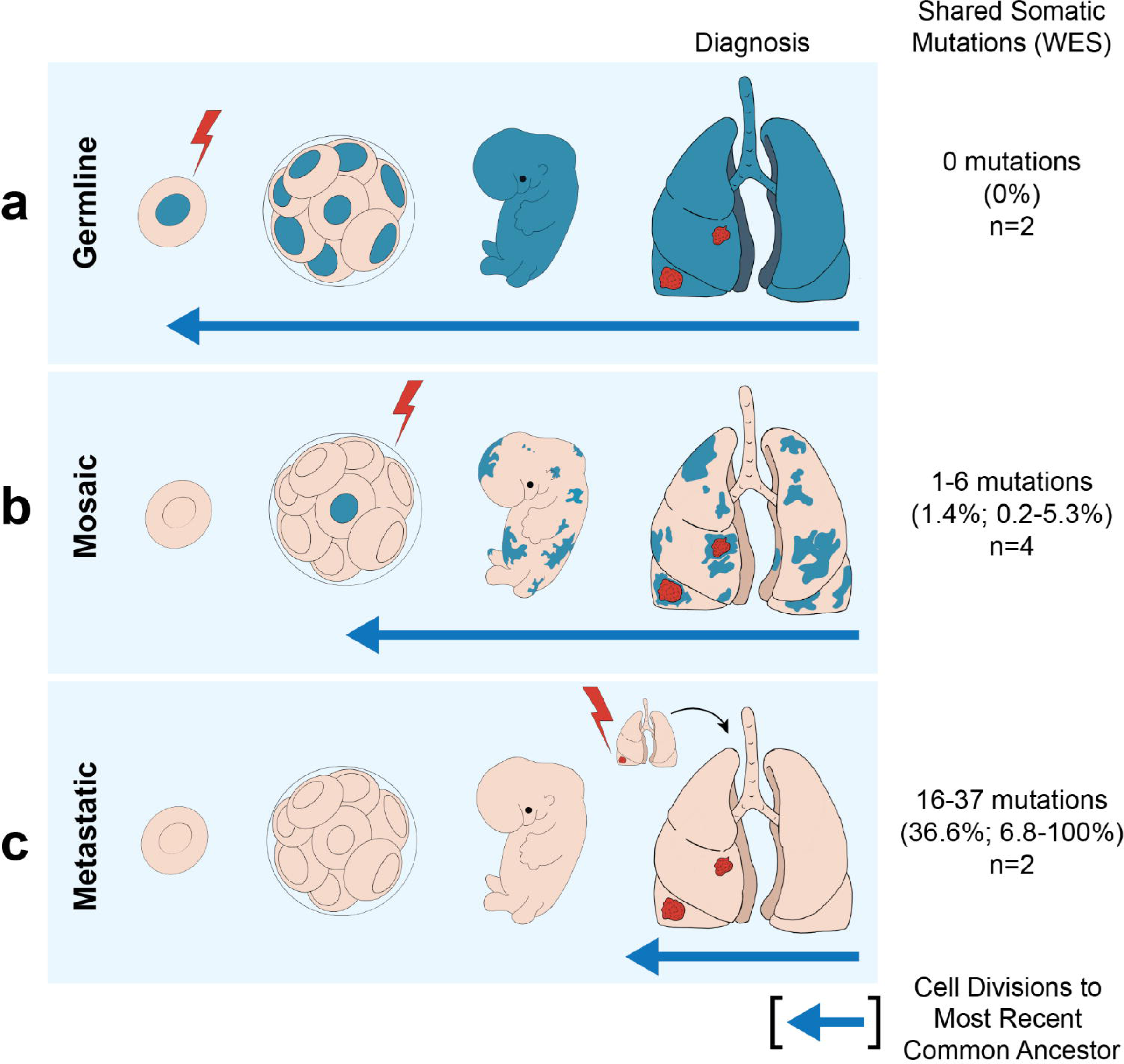
Genetic distinctions between multiple lung cancers with inherited, mosaic and metastatic origin. Schematic representation of three distinct mechanisms underlying multiple *EGFR*-mutant lung tumors. The average percent of somatic mutations shared among different tumors is shown on the right (range in parentheses). **a,** In cases with inheritance of an attenuated mutant *EGFR* allele (either familial or apparent sporadic), the mutation is present in the germline (lightning bolt) and present in all somatic tissues. A second canonical *EGFR* mutation arises somatically at high frequency in predisposed lung cells, leading to multiple tumors with no shared somatic mutations. The evolutionary distance to the most common shared ancestor between different tumors (arrow) extends to the germline. 0 non-germline mutations are shared between tumors. **b,** Cases with mosaic predisposition arise from acquisition of an *EGFR* mutation during development (lightning bolt). This timing determines the proportion of normal cells containing the variant allele and the likelihood of developing multiple tumors. In addition to the activating *EGFR* mutation, a small number of additional somatic genetic variants are shared before the mosaic tissues diverge and acquire independent tumor-associated mutations. **c,** In sporadic cancers without genetic susceptibility, a single *EGFR* mutation arises in a somatic lung epithelial cell (lightning bolt), generating a single tumor. Metastases from this tumor share extensive mutational profiles and the most recent common ancestor for these multiple tumors is the primary tumor.

The presence of multiple distinct primary *EGFR*-mutant cancers at the time of clinical presentation has long presented a conundrum. Two common hypotheses include intrapulmonary metastasis and field cancerization. Lung cancer metastasis characteristically presents with disseminated disease, but it may also arise through intrapulmonary lymphatic spread within adjacent regions of the lung. Local spread through air spaces (STAS) has also been reported, particularly in some tumors with micropapillary and/or solid histology^48^. The detection of shared mutations between anatomically distinct primary lung tumors has made intrapulmonary spread the lead model to explain the presence of multiple primary tumors at presentation. Challenging this model of localized metastasis, however, is the fact that many lesions in patients with multiple primaries are histologically classified as pre-invasive, and even pre-neoplastic. Furthermore, their anatomical locations may include different lobes or contralateral lungs, which are likely beyond the reach of localized lymphatic or airspace spread. In the cases reported here, we did observe two cases with metastatic disease, as suspected clinically and confirmed by their largely shared mutational composition. However, the four cases in which anatomically distinct primaries contain a number of shared somatic mutations that comprise only a small fraction of the total exonic mutational burden are inconsistent with intrapulmonary metastasis.

The second common hypothesis, “field cancerization”, is a phenomenon whereby the entire tissue is damaged by carcinogenic exposure leading to multiple independent and genetically unrelated tumors. It is most frequently applied to cases with a heavy smoking history, although exposures due to environmental radon and occupational exposures have also been considered. Recent studies in barrier organs, including skin and esophagus, have indicated that as they age, histologically normal tissues may acquire canonically oncogenic mutations that give rise to small patches of clonal expansion, but without evidence of frank malignancy^30–34^. In the lung, deep sequencing reveals that up to 18% of normal samples have a detectable mutant *EGFR* allele, and even more cases harbor *KRAS* and other mutated oncogenes^29^. The high prevalence of normal lung specimens with rare detectable *EGFR*-mutant alleles due to field cancerization does not explain the occurrence of multiple *EGFR*-mutant primary tumors described here, since they would be genetically independent, lacking the multiple shared mutations that define the developmental mosaically-derived primaries.

We propose that a canonical and fully activating *EGFR* mutation may arise early during lung development, creating a mosaic of lung epithelial cells harboring this mutation and distributed across the adult organ. Our analysis of poly-G repeats in such tumors suggests that *EGFR* mutations may occur before cells have undergone half of the divisions on their way to tumor initiation. While normal tissues other than lung were not available for our retrospective cohort, we were able to show the presence of a low frequency of *EGFR* mutations in normal lung tissues, only in the patients with multiple primaries whose tumors had the corresponding *EGFR* mutation. Unlike mutant *EGFR* alleles in the context of field cancerization, which represent random independent genetic events, these *EGFR* mutations within normal tissues may result from early developmental mosaicism, ultimately giving rise to anatomically disparate tumors that also share multiple passenger mutations, consistent with their common clonal origin. The mechanism by which *EGFR*-mutant lung epithelial cells generate pre-malignant and ultimately invasive cancers is unknown, but recent epidemiological and mouse modeling studies indicate that inflammation associated with air pollution enhances the likelihood that an *EGFR*-mutant cell will give rise to a malignant tumor^29^. In this context, the high frequency of mutant *EGFR* allele detection in normal lung is in marked contrast to their virtual absence (<0.1%) in normal skin^29^, raising the possibility that EGF signaling mediates distinct proliferative effects in the lung, thereby contributing to the persistence of mosaically-derived progeny during early lung development.

The concept of mosaicism in human genetics is best illustrated by Neurofibromatosis type 1, where up to 5% of patients have segmental café au lait spots, attributable to a mutation in the *NF1* gene that arose during embryonic development and affects only a portion of the adult body^49,50^. In the brain, post-zygotic mosaicism affecting Ras/Raf/MAPK signaling may play a role in the pathogenesis of mesial temporal lobe epilepsy^51^. In cancer, the concept of a clonally-derived origin for multifocal sporadic cancers was first proposed based on X chromosome inactivation studies in bladder cancer^52^. Mosaic inactivating mutations have been reported in tumor suppressor genes linked to high-risk cancer predisposition syndromes, including Li-Fraumeni and von Hippel Lindau syndromes^53,54^. In the pediatric kidney cancer Wilms tumor, geographically distinct precursor lesions, called nephrogenic rests, share mutations with each other and with geographically distinct tumors, consistent with mutations arising during early renal organogenesis^42^. While developmental mosaicism has not been reported as a common event in epithelial cancers affecting adults, one case report described the incidental discovery at autopsy of multiple precancerous lung lesions, all sharing an identical somatic *TP53* mutation raising the possibility of a mosaic mechanism^55^.

The second genetic mechanism that we describe underlying multiple *EGFR*-mutant primary lung cancers extends the observation of rare familial lung cancer to cases without such pedigrees. As with familial transmission of a germline T790M-*EGFR* allele, the germline *EGFR* variants identified here encode mutant proteins with modestly increased enzymatic activity, suggesting that, unlike strongly activating mutants, they are not deleterious during normal embryonic development. They may, however, be sufficient to increase the size of target cell populations within the lung, which undergo tumorigenesis after sustaining a second more strongly activating *EGFR* mutation. To date, clinical sequencing efforts of *EGFR* in NSCLC have identified approximately 500 somatic variants of unknown significance, a small subset of which have been identified in the germline (ClinVar and gnomAD) ^36,56,57^. Further studies will be required to determine how many additional *EGFR* variants have subtly enhanced signaling properties when present in the germline, potentially linked to increased tumorigenesis, and their penetrance.

Finally, we note that understanding the etiology of multiple *EGFR*-mutant tumors with synchronous presentation may impact clinical treatment strategies. Management of independent primary tumors includes lung parenchyma-sparing resections with curative intent, and mosaically-derived tumors should not be confused with metastatic disease, despite the presence of some shared mutations. Moreover, when due to either a germline *EGFR* variant or to developmental mosaicism, patients presenting with multiple *EGFR*-mutant primary tumors are likely at risk for developing additional tumors during their lifetime, suggesting the importance of ongoing surveillance and raising consideration of novel prophylaxis strategies.

## Methods

### Patients

This study was approved by the Institutional Review Board of Massachusetts General Hospital and complies with all relevant ethical regulations. Ten patients were selected retrospectively from an institutional database as having undergone resection for two or more early-stage lung cancers, with at least one lesion having a known EGFR mutation by NGS sequencing analysis. These patients gave informed consent for their biological materials to be included in this database. These tumors were classified as early stage by the reviewing pathologist at the time, even though our genetic analysis suggests that two of the patients have metastatically related tumors. No patients had received systemic treatment for cancer preoperatively. Clinical histories were extensively reviewed (See Table 1 and Supplementary Table 1 for summary of patient and tumor characteristics) Patient genetic ancestry was inferred from electronic health records. An additional two patients with a known inherited EGFR T790M mutation were included for proof-of-concept.

### Specimen collection and histopathology

Sections from the entire tumor and representative lung parenchyma distinct from the tumor were fixed in formalin and embedded in paraffin. Histopathologic slides were made from the formalin fixed paraffin embedded tissue blocks (FFPE) and retrospectively reviewed by a single observer, expert pulmonary pathologist (M.M.K) who histologically classified and staged individual tumors in accordance with WHO classification of lung tumors^58^ and the 8^th^ Edition American Joint Committee on Cancer (AJCC) lung cancer staging guidelines, respectively. The area of highest tumor purity and histologically normal lung tissue were also selected.

### TNA extraction protocol

Total nucleic acid was extracted from designated tumor and normal lung tissue from each patient using the standard protocol of Agencourt Formapure kit (Supplementary Table 1). The approximate location of the normal samples relative to the tumors is shown in Supplementary Figure 1A.

### Whole exome sequencing

Prior to whole-exome sequencing (WES) a standardized PicoGreen® dsDNA Quantitation Reagent test (Invitrogen, Carlsbad, CA) was used to quantify DNA in triplicate. The Fluidigm Genotyping fingerprint genotyping of 95 frequent SNPs was used for the quality control identification check (Fluidigm, San Francisco, CA). Using the KAPA Library Prep kit and palindromic forked adapters from Integrated DNA Technologies, libraries were constructed from double-stranded DNA. Prior to hybridization, libraries were combined. Utilizing a 37Mb target, hybridization and capture were carried out using the essential components of Illumina’s Rapid Capture Enrichment Kit. On the Agilent Bravo liquid handling system, the library building, hybridization, and capture processes were all fully automated. Library pools were denatured on the Hamilton Starlet using 0.1N NaOH following post-capture enrichment. DNA libraries were cluster amplified using HiSeq 4000 exclusion amplification reagents and HiSeq 4000 flowcells in accordance with the manufacturer’s (Illumina) instructions. HiSeq 4000 flowcells were sequenced using Sequencing-by-Synthesis chemistry. RTA v.2.7.3 or later was then used to examine the flowcells. Sequencing of each pool of entire exome libraries was done using paired 76 cycle runs with two 8 cycle index reads across the number of lanes required to provide coverage for all libraries in the pool.

Sequence data were analyzed using the Broad Institute’s Cancer Genome Analysis WES Characterization Pipeline, in which aligned BAM files were inputted into a standardized WES somatic variant-calling pipeline as previously described^59^ that included MuTect for calling somatic single nucleotide variants (sSNVs), Strelka2 for calling small insertions and deletions (indels), deTiN for estimating tumor-in-normal (TiN) contamination, ContEst for estimating cross-patient contamination, AllelicCapSeg for calling allelic copy number variants, and ABSOLUTE for estimating tumor purity, ploidy, cancer cell fractions, and absolute allelic copy number. Artifactual variants were filtered out using a token panel-of-normals (PoN) filter, a blat filter, and an oxoG filter. The PhylogicNDT^59^ suite of tools was used to generate posterior distributions on cluster cancer cell fractions and mutation membership to calculate the ensemble of possible trees that support the phylogenetic relationship of detected cell populations. Through applying this tool across a set of samples for the patient, the most likely tree was identified if samples were clonally related and independent phylogenies were delineated for unrelated samples.

### Polyguanine genotype data pre-processing

Generation and analysis of polyguanine repeat (poly-G) genotypes was performed as previously described^44–46^. Briefly, 33 poly-G loci were PCR-amplified using primers targeting their flanking sequences. All reactions were run in duplicate. PCR product length was measured using an ABI 3730xl DNA Analyzer and exported as tab-delimited text files through the ThermoFisher Microsatellite Analysis Tool (https://www.thermofisher.com/us/en/home/cloud/all-analysis-modules/sanger-analysis-modules.html). Reactions whose intensities were less than 10% of the average intensity for that patient and locus were excluded. If the length distributions of both duplicates were similar (Jensen-Shannon Divergence < 0.11), the duplicate with higher fluorescence intensity was picked as the representative replicate. At a larger discordance between length distributions, the poly-G tract was excluded from analysis in all samples of that patient. More details on filtering and quality control of poly-G genotypes are provided in Naxerova et al, 2017 (Ref. ^45^).

### Polyguanine genotype data analysis

Amplification of microsatellites produces a characteristic fragment stutter pattern due to polymerase slippage during PCR. The mean fragment length at each locus, which represents the genotype of the most recent common ancestor of all sampled cells^60^ was used to simplify this stutter pattern to a single value. Somatic shifts in poly-G length (mutations) are reported in relation to the normal (germline) sample from each patient. In order to construct phylogenetic trees using the mean length of poly-G markers, distance matrices containing all the samples from one patient were constructed using the Manhattan distance (preprint forthcoming). This distance measures the sum of insertions and deletions among all poly-G markers in two samples, normalized by the number of poly-G markers analyzed. Because the Manhattan distance simply counts the number of mutations, it scales linearly with the number of cell divisions separating two samples. However, the Manhattan distance is affected by a sample’s purity because the presence of normal cells within a tumor reduces the mean length. Based on the distance matrices, phylogenetic trees were constructed using the neighbor-joining method implemented in the R package ape^61^. Evolutionary distance between two tumors was estimated using Pearson’s correlation coefficient (r) between the two vectors of poly-G marker lengths. Only patients in which at least half of all poly-G markers could be successfully amplified across all samples were considered for this analysis. Samples were analyzed in two batches and only tumors that were analyzed in the same batch were compared to each other. Pearson’s correlation coefficient estimates the fraction of cell divisions in the history of two tumors that they have spent as part of the same lineage. A correlation of 0 means the lineages giving rise to two tumors split at the zygote stage and that they share 0% of their cell divisions, while a correlation of 1 means that the tumors’ lineages coincide and that they share 100% of their cell divisions. Pearson’s correlation coefficient only compares the direction of mutations, but not their magnitude, thus its estimation of evolutionary distance is not affected by purity and mutation rate. To assess the expected correlation value of two unrelated tumors, the distribution of r was calculated based on tumors from different patients, across all possible tumor pairs in this cohort in which at least 15 of the same poly-G loci were successfully amplified in both samples.

### EGFR mutant construct

The WT EGFR expression plasmid pHAGE-EGFR was a gift from Gordon Mills & Kenneth Scott (Addgene plasmid #116731) ^62^. EGFR mutant constructs containing patient-specific DNA mutations were generated by site-directed mutagenesis (Agilent QuikChange® II XL) and confirmed by Sanger sequencing. Overall expression of ectopic EGFR constructs was approximately 10-fold higher than the level of endogenous expression in human NIH2228 cells, a lung cancer cell line that expresses moderate levels of EGFR. These constructs are available upon request.

### Immunoblot analysis

Cells were lysed with RIPA buffer (Sigma Aldrich # R0278) containing protease and phosphatase inhibitors (Life Technologies #A32965; A32957). Lysate was cleared and immunoblotted according to standard protocols. Antibodies used are as follows: EGFR (Cell Signaling Technologies #4267 1:500 dilution in 5% BSA in PBST, imaged on LiCor); EGFR pY845 (Cell Signaling Technologies #6963 1:500 dilution in 5% BSA in PBST, imaged on LiCor); Akt1/2 (Cell Signaling Technologies #9272 1:1000 dilution in 5% milk in PBST, imaged on film); Akt pS473 (Cell Signaling Technologies #4060 1:500 dilution in 5% milk in PBST, imaged on film); Vinculin (Sigma Aldrich MAB3574 1:2000 dilution in 5% BSA in PBST, imaged on LiCor).

### Transformation assays

For soft agar colony formation assays, NIH/3T3 cells stably expressing RFP, WT EGFR, or mutant EGFR were suspended in DMEM + 10% FBS containing 0.4% agarose with no additional EGF for three weeks. Colony growth was assayed by staining with 0.2% crystal violet in methanol for 10 min, followed by manual counting.

### Droplet Digital PCR analysis

ddPCR to detect EGFR L858R mutations was performed on total nucleic acid extracted from tissue slides. We used the commercially validated probe-set for EGFR WT and p.L858R c.2573T>G (BioRad Assay ID dHsaCP2000022). Samples were prepared following standard protocol (Biorad). Briefly, 2x ddPCR Supermix for probes (no dUTP) was combined with 20-400 ng patient DNA, 1x primer/probe mix, and 5U MseI restriction enzyme (NEB). After droplet generation, samples were thermocycled with an annealing/extension temperature of 55°C.

## Disclosures

D.H. is supported by the Howard Hughes Medical Institute (HHMI) and the National Institutes of Health (5R01CA137008). L.V.S. is supported by the National Institutes of Health (1P50CA265826, R37CA225655), the Lungstrong Foundation and The Landry Family. L.V.S. has institutional research funding from AstraZeneca, Novartis and Delfi Diagnostics and has received consulting fees from AstraZeneca, Janssen, Pfizer and Genentech. I.L. serves as a consultant for PACT Pharma Inc. and has stock, is on the board and serves as a consultant for ennov1 LLC., is on the board and holds equity in Nord Bio, Inc. G.G is supported by the Broad-IBM cancer resistance project and is the Paul C. Zamecnik Chair in Oncology at the Massachusetts General Hospital Cancer Center. G.G. receives research funds from IBM and Pharmacyclics, and is an inventor on patent applications related to MSMuTect, MSMutSig, MSIDetect, POLYSOLVER, SignatureAnalyzer-GPU and MinimuMM-seq. G.G. is a founder, consultant and holds privately held equity in Scorpion Therapeutics.

## Data Availability

All data produced in the present study are available upon reasonable request to the authors.

## Data availability statement

WES data will be deposited at dbGaP upon publication. WES data are currently available from the authors by reasonable request.

## Code availability statement

The code used to analyze the poly-G genotypes will be deposited at GitHub upon publication. Code is currently available from the authors by reasonable request.

## Supplementary Tables (Excel files)

**Supplementary Table 1. Clinical characteristics of individual tumors from each patient.** Tumors and normal samples are listed, with the normal tissues listed next to the matched resected tumor. Tumor size represents the greatest dimension.

**Supplementary Table 2. Whole Exome Sequencing results for multiple primary lung cancers.** For each patient, the cancer cell fraction (ccf) of any mutation that was found in at least one sample is reported for every sample.

**Supplementary Table 3. Summary of WES and poly-G data.** For each tumor across individual patients, the number of shared and total mutations identified by WES is noted, along with the fraction of cell divisions shared according to the poly-G data analysis.

## Supplementary Figure legends

**Supplementary Figure 1.**
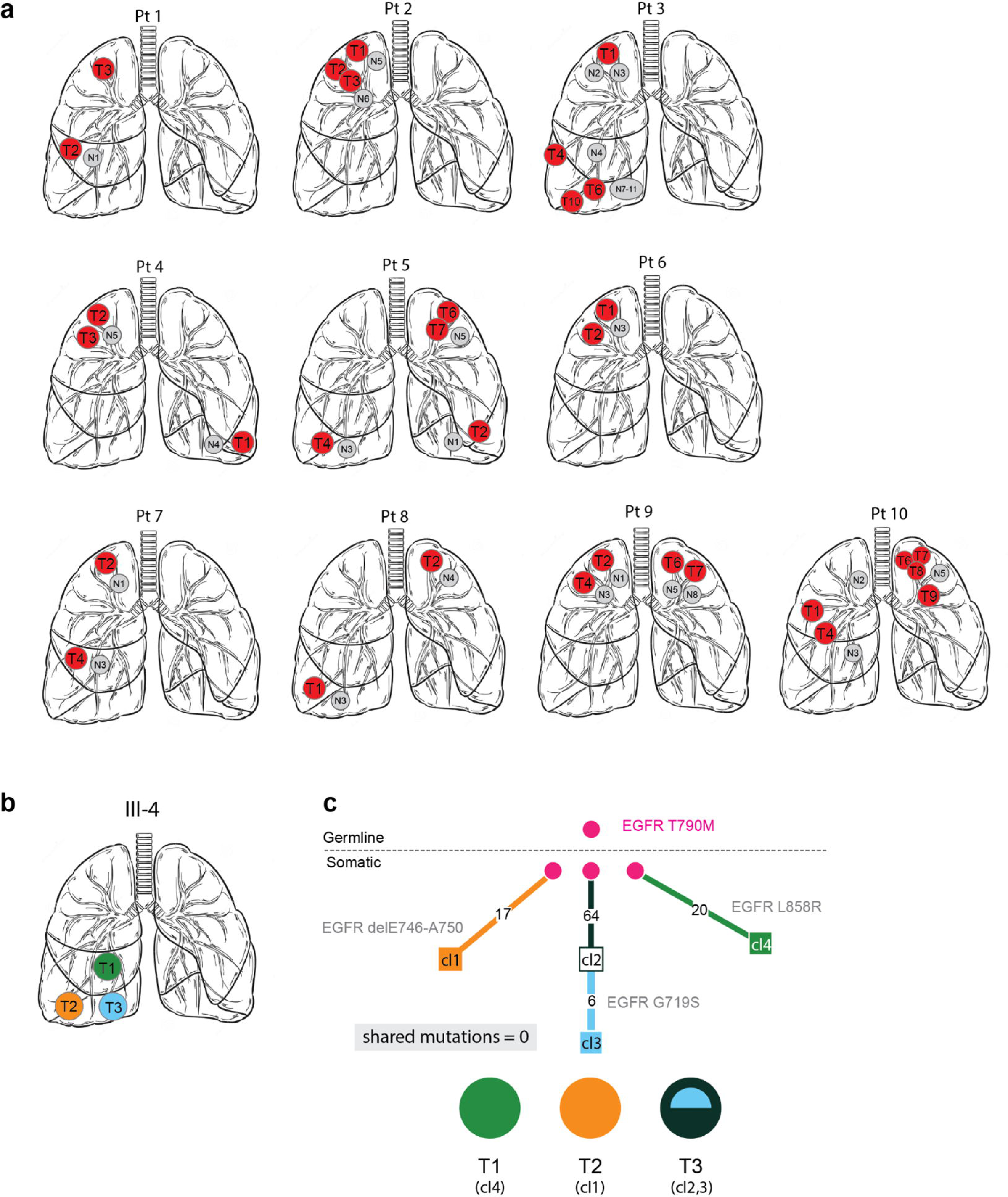
**a,** Schematics of the relative location of all normal samples and corresponding tumors in our cohort. **b,** Schematic of the tumor locations in patient III-4. **c,** Lineage tracing of patient III-4 with a known germline *T790M-EGFR* mutation, identified by Whole Exome Sequencing. The germline *EGFR* mutation found in normal lung tissue is denoted at the top of the tree. Numbers on branches are mutations that accumulated between two nodes, with known cancer-associated *EGFR* mutations highlighted next to the branch.

**Supplementary Figure 2.**
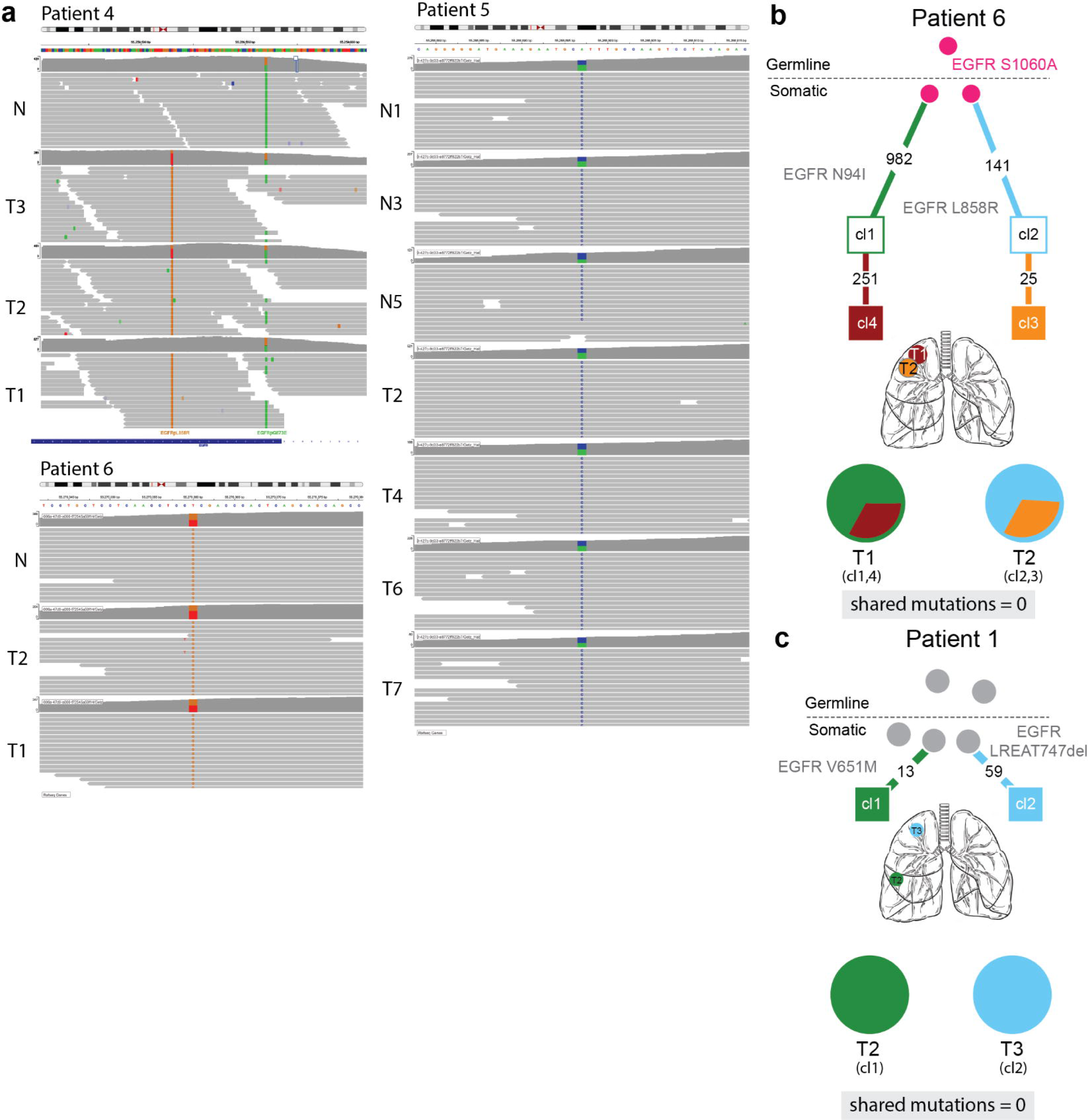
**a,** IGV tracks for the patients harboring germline variants, showing the germline *EGFR* mutations in the normal samples as well as tumors. Patient 4 also shows *EGFR G873E* on same read as *L858R* in the tumor samples, indicating that the two mutations are *in cis*. **b,** Phylogenetic tree from patient 6, with a germline variant identified by WES. The germline *EGFR* mutation that is shared among tumors from the same individual is shown in magenta. Numbers on branches are mutations that accumulated between two nodes. **c,** Phylogenetic tree from patient predicted to be independent by WES. As observed in patients with a germline *EGFR* mutation, there is no intersection between the branches, but unlike these patients, there is no germline mutation shown in magenta.

**Supplementary Figure 3.**
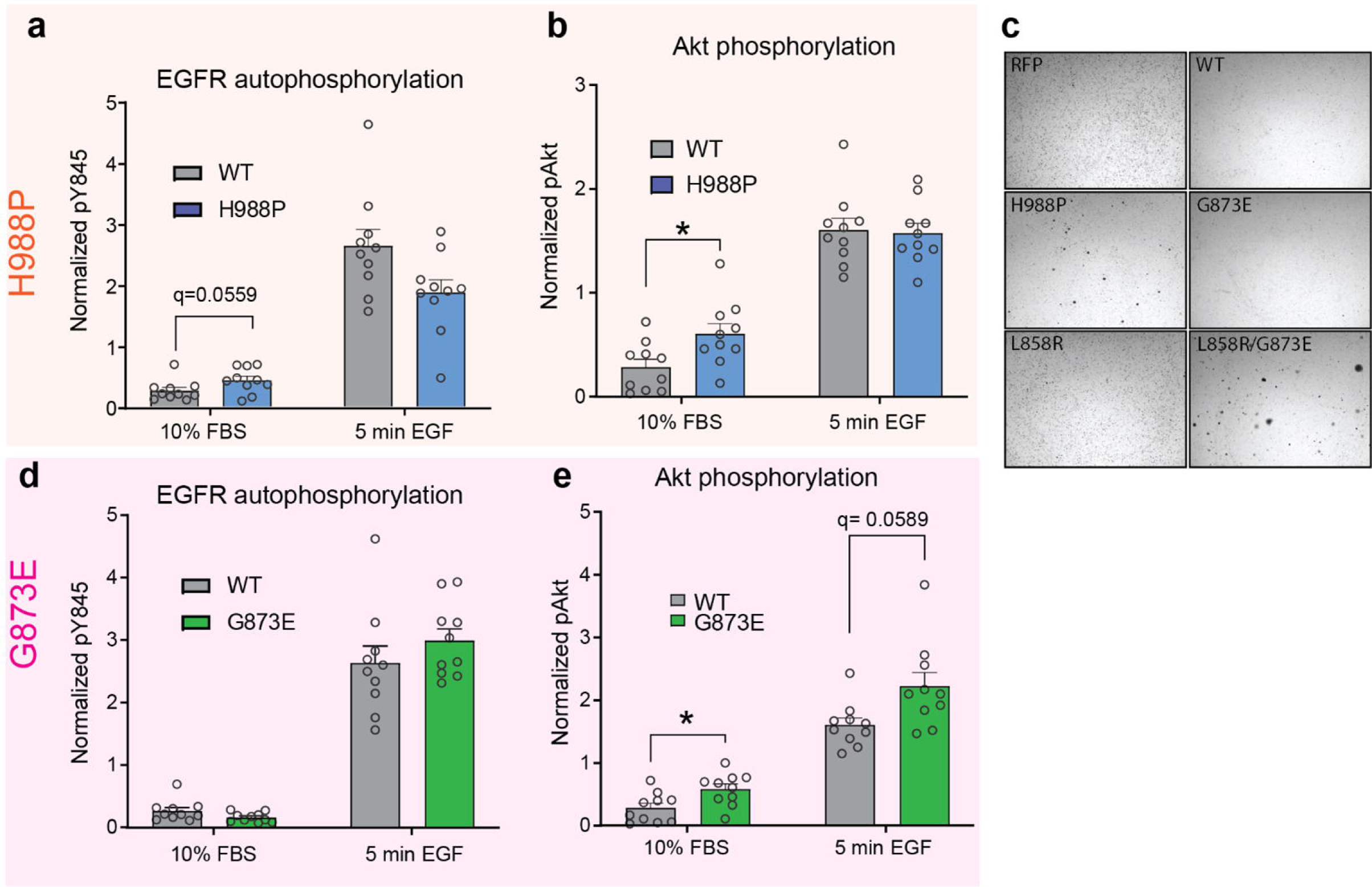
**a-b,d-e** Quantification of ten replicates of Figure 2d-e, g-h. EGFR autophosphorylation at Y845 was normalized to vinculin loading control, then phosphorylation was normalized to total EGFR expression in that sample, and finally normalized to the average signal within an experiment for comparison across experiments. AKT phosphorylation at S473 was normalized to vinculin loading control, and then to the average signal within an experiment. Error bars are SEM. False Discovery Rate q-values were corrected for multiple comparisons by the Benjamini, Krieger and Yekutieli procedure. * q<0.05 **c,** Representative images of a soft agar colony formation assay corresponding to Figure 2f.

## Notes

### Funding Statement

This study was funded by the Howard Hughes Medical Institute and IBM Research.

### Author Declarations

IRB of Massachusetts General Hospital gave ethical approval for this work.

